# Evaluating the causal impact of reproductive factors on breast cancer risk: a multivariable mendelian randomization approach

**DOI:** 10.1101/2024.02.02.24301815

**Authors:** Claire Prince, Laura D Howe, Eleanor Sanderson, Gemma C Sharp, Abigail Fraser, Bethan Lloyd-Lewis, Rebecca C Richmond

## Abstract

**Background:** Observational evidence proposes a protective effect of having children and an early age at first birth on the development of breast cancer, however the causality of this association remains uncertain. In this study we assess whether these reproductive factors impact breast cancer risk independently of age at menarche, age at menopause, adiposity measures and other reproductive factors that have been identified as being causally related to or genetically correlated with the reproductive factors of interest.

**Methods:** We used genetic data from UK Biobank (273,238 women) for reproductive factors, age at menarche and menopause, and adiposity measures, and the Breast Cancer Association Consortium for risk of overall, estrogen receptor (ER) positive and negative breast cancer as well as breast cancer subtypes. We applied univariable and multivariable Mendelian randomization (MR) to estimate direct effects of ever parous status, ages at first birth and last birth, and number of births on breast cancer risk.

**Results:** We found limited evidence of an effect of age at first birth on overall or ER positive breast cancer risk in either the univariable or multivariable analyses. While the univariable analysis revealed an effect of later age at first birth decreasing ER negative breast cancer risk (Odds ratio (OR): 0.76, 95% confidence interval:0.61-0.95 per standard deviation (SD) increase in age at first birth), this effect attenuated with separate adjustment for age at menarche and menopause (e.g., OR 0.83, 0.62-1.06 per SD increase in age at first birth, adjusted for age at menarche). In addition, we found evidence for an effect of later age at first birth on decreased human epidermal growth factor receptor 2 enriched breast cancer risk but only with adjustment for number of births (OR 0.28 (0.11-0.57) per SD increase in age at first birth).

We found little evidence for direct effects of ever parous status, age at last birth or number of births on breast cancer risk, however, analyses of ever parous status and age at last birth were limited by weak instruments in the multivariable analysis.

**Conclusions:** This study found minimal evidence of a protective effect of earlier age at first birth on breast cancer risk, while identifying some evidence for an adverse effect on ER negative breast cancer risk. However, multivariable MR of ever parous status and age at last birth is limited by weak instruments which might be improved in future studies with larger sample sizes and when additional genetic variants related to reproductive factors are identified.

## Introduction

Reproductive factors relating to pregnancy, including ever having children, the number of children a woman has and age at first and last birth, have been found to be related to the development of breast cancer.^(1–3)^ However, the relationships between these factors and breast cancer risk are complex, and the underlying mechanisms remain unclear.

Experiencing pregnancy earlier in life has been shown to be associated with a reduced long-term breast cancer risk in previous studies,^(2,4–8)^ with one study identifying a 7% increase in risk (odds ratio (OR): 1.07; 95% confidence interval (CI): 1.01-1.13) per 5-year increase in age at first full-term pregnancy.^(5)^

There are several proposed mechanisms which may underlie the protective role of an early pregnancy on breast cancer development. Parity stimulates the differentiation of mammary epithelial lobules to its mature state, and cells in a differentiated state have reduced susceptibility to mutation and tumorigenesis.^(9,10)^ Additionally parous, compared to nulliparous, women have lower responsiveness to estrogen, via a downregulation of estrogen receptor (ER) α, and upregulation of ERβ.^(11)^ ERα have higher affinity to estrogen compared to ERβ, and heterodimerising of ERα and ERβ reduces the affinity of ERα to estrogen.^(12)^ Since estrogen is known to be proliferative and genotoxic, there may be an increased risk of mutation and subsequent development of breast cancer among nulliparous women.^(9,13)^

However, a short-term adverse effect of pregnancy on breast cancer risk has also been observed (for women aged 35 years at first birth, 5 years post-delivery OR of 1.26; 1.10-1.44, compared to nulliparous women).^(14)^ This short-term elevated risk could occur mainly through two mechanisms.^(5)^ Firstly, levels of ovarian hormones, which have proliferative effects, increase during pregnancy and subsequently there is a higher chance of a mutation occurring.^(9)^ Secondly, involution, that occurs following lactation, involves programmed cell death and tissue remodelling of the breast which has similar properties to wound healing and inflammation,^(13,15,16)^ which are known to be pro-oncogenic.^(13,15)^

The long-term protective benefits of pregnancy may therefore not emerge until at least 10 years after delivery,^(5,13–15)^ with a proposed cross-over point where the group at higher risk changes from parous women to nulliparous women.^(14,15,17–19)^

Considering the proposed mechanism linking age at first pregnancy and breast cancer development, it is thought that a shorter interval between menarche and first pregnancy reduces breast cancer risk given the prevalence of undifferentiated cells that are more susceptible to mutation-causing events during this time.^(13)^ The first pregnancy is therefore mainly protective against later breast cancer risk when experienced earlier in life. In addition, a late age at first pregnancy may increase risk because the proliferative effects of pregnancy cause underlying mutations, that arise during the high-risk window, to be exacerbated and result in cancer formation.^(4,20)^ There appears to be little difference in risk if the first pregnancy occurs between 30 and mid-30s compared to nulliparous women, while having the first pregnancy after the age of 35 may increase risk.^(2,4)^ However, evidence for this increased risk is not consistent across all studies.^(2,21,22)^

Considering the role of multiparity, there is evidence to suggest that each additional pregnancy conveys further protection beyond an early age at first pregnancy.^(16,23)^ While other studies show that a higher age at last birth is associated with higher breast cancer risk,^(7,8,24–26)^ with one study reporting an increased breast cancer risk per every 5-year rise in age at last birth (OR: 1.09; 1.04-1.13).^(25)^ However, the mechanisms by which number of births and age at last birth lead to breast cancer are unclear.^(7,27)^

Observational studies are known to be limited by confounding bias as it is difficult to capture all confounders accurately. Mendelian randomisation (MR), a method less likely to be affected by confounding and reverse causation,^(28,29)^ has been used to assess the causal relationship between reproductive factors and breast cancer risk by using genetic variants robustly associated with the exposure as instruments. MR studies have consistently found little evidence of an effect of age at first birth,^(30,31)^ (OR 0.92; 0.79-1.07^(30)^), ^(30)^but interestingly, some evidence for a protective effect of a later age at last birth on breast cancer risk (OR 0.69; 0.54-0.88), in contrast to observational data.^(31)^ An MR identified an estimate reflecting a higher number of births reduces breast cancer risk although confidence intervals span the null (OR 0.70; 0.44-1.11).^(30)^

It is currently unclear how each reproductive event affects risk in isolation since these traits are highly correlated with, and/or causally linked to other reproductive factors as well as age at menarche and menopause, and adiposity measures,^(32,33)^ which are established breast cancer risk factors.^(34,35)^ Multivariable MR (MVMR) is an extension of MR which can estimate the direct effect of two or more risk factors while accounting for genetic correlation between them,^(36,37)^ which may be valuable for investigating the impact of correlated reproductive factors.

The effects of reproductive factors on risk of different subtypes of breast cancer are not well understood.^(1,30,38–44)^ There is some consensus that a lower age at first birth and higher parity decreases risk of hormone-receptor positive breast cancer subtypes, however there is less consistency for the effects of these traits on risk of hormone-receptor negative subtypes.^(1,41–44)^ Hormone-receptor negative subtypes are more common in women under 40,^(45)^ therefore, pregnancy may increase short-term risk for these subtypes more so than those classified as hormone-receptor positive. Additionally, as more women are choosing not to have children,^(46)^ or having children later in life,^(47)^ it is important to understand how these traits might convey different risk depending on subtype.

### Aims

We aimed to use univariable and multivariable MR methods to untangle the effects of ever having children, age at first birth, age at last birth and number of children birthed on breast cancer risk. We further aimed to investigate whether effects are independent of age at menarche and menopause, and adiposity measures as well as other reproductive factors, and whether effects differ for ER positive compared to ER negative breast cancer.

Furthermore, based on these findings we aimed to assess the effects of reproductive factors on risk of intrinsic breast cancer subtypes.

## Methods

### UK Biobank

The UK Biobank study is a large population-based cohort of 502,682 individuals who were recruited at ages 37–73 years across the UK between 2006 and 2010. The study includes extensive health and lifestyle questionnaire data, physical measures, and biological samples from which genetic data has been generated. The study protocol is available online, and more details have been published elsewhere.^(48)^ At recruitment the participants gave informed consent to participate and be followed up. Unless stated otherwise, in UK Biobank, the factors investigated in the current study were derived from questionnaire responses at the baseline assessment.

#### Reproductive factors

The reproductive factors investigated in this study were: age at first live birth, age at last live birth, number of live births and parous status (ever/never given birth at the time of assessment). Hereafter these will be referred to as age at first birth, age at last birth, number of births and ever parous status. In UK Biobank these reproductive factors were derived from questionnaire responses at the baseline assessment, further details are in **Supplementary File 1**.

To identify genetic variants robustly related to each of the reproductive factors, we performed genome-wide association study (GWAS) for each reproductive factor among women in the UK Biobank. Each GWAS was performed using the Medical Research Council (MRC) Integrative Epidemiology Unit (IEU) UK Biobank GWAS pipeline,^(49,50)^ and further details can be found in **Supplementary File 1.**

#### Additional variables

We additionally included age at menarche and menopause, and adiposity measures in childhood and adulthood. Further details on how age at menarche and menopause were defined can be found in **Supplementary File 1**. We explored adiposity in childhood using the comparative body size measure obtained from the baseline questionnaire in UK Biobank. Participants were asked, “When you were 10 years old, compared to average would you describe yourself as:”, and were given the options: “Thinner”, “Plumper” and “About average”. We investigated adiposity in adulthood using body size based on BMI. BMI was derived from height and weight measured during the initial UK Biobank Assessment Centre visit. The categorical body size measure was composed on three groups based on the same proportions as the childhood body size variable.

We performed GWAS for age at menarche and menopause among women in the UK Biobank similarly to the reproductive factors stated above. We obtained female only GWAS summary statistics for childhood and adulthood body size from Richardson et al. 2020, where they performed GWAS using a similar approach.^(51)^

### Breast cancer association consortium

#### Overall breast cancer

The overall breast cancer risk GWAS summary statistics were obtained from the Breast Cancer Association Consortium (BCAC) which used iCOGS, OncoArray and other GWAS data for 133,384 breast cancer cases and 113,789 controls of European ancestry from 82 studies.^(52)^

#### Estrogen receptor status

GWAS summary statistics for breast cancer risk stratified by ER status were obtained from BCAC.^(53)^ The data contained 69,501 ER positive cases, 21,468 ER negative cases and 105,974 controls of European ancestry.

#### Estrogen receptor negative subtypes

We additionally investigated the birth-related reproductive factors, which showed an effect on risk of ER positive or negative breast cancer, in relation to intrinsic breast cancer subtypes that are characterised by being hormone-receptor positive or negative. GWAS summary statistics used in this study were for risk of two breast cancer subtypes; human epidermal growth factor receptor 2 (HER2) enriched (718 cases) and triple negative (2,006 cases), were obtained from the BCAC. The data involved individuals of European ancestry and contained 20,815 controls.^(52)^

### Genetic correlation

Genetic correlations between the reproductive factors, age at menarche and menopause, adiposity measures and breast cancer risk outcomes were calculated using linkage disequilibrium score regression (LDSC) and the UK Biobank, and BCAC GWAS summary statistics.^(54,55)^ Further details can be found in **Supplementary File 1**.

### STROBE-MR

The Strengthening the Reporting of Observational Studies in Epidemiology using Mendelian Randomization (STROBE-MR) guidelines have been followed in the analysis of this work (**Supplementary File 1**).^(56,57)^

### Univariable analysis

We performed univariable MR (UVMR) of ever parous status, age at first birth, age at last birth and number of births on overall, ER positive and ER negative breast cancer risk. We additionally performed UVMR on reproductive factors that appear to have an effect on risk of ER negative breast cancer, on ER negative breast cancer subtypes (HER2 enriched and triple negative). We used the function “mr()” from TwoSampleMR R package.^(50)^ The primary analysis focused on the inverse variance weighted (IVW) MR method.^(58)^ GWAS estimates for age at first birth, age at last birth, number of births were standardized based on the standard deviation of the phenotypic exposure (mean = 0 and standard deviation (SD) = 1) prior to performing MR.

#### Evaluating univariable mendelian randomization assumptions

Mendelian randomization has three main assumptions: the relevance assumption states the genetic variants instrumented are associated with the exposure of interest; the independence assumption states the genetic variants and outcome under investigation do not share any common causes; the exclusion restriction assumption states that the instrumented genetic variants do not affect the outcomes via any pathway other than the exposure being examined.^(59)^

To evaluate the strength of the genetic instruments, we determined the mean F statistic for each trait, ^(59)^ and considered a F statistic of 10 or above as indicative of a strong instrument. To evaluate whether the genetic instruments are pleiotropic, we performed MR using additional methods: Weighted mode, ^(60)^ Weighted median,^(61)^ and MR Egger. ^(62,63)^ The intercept from MR Egger regression was used to determine directional pleiotropy.^(62)^

We also applied MR-PRESSO (Mendelian Randomisation Pleiotropy RESidual Sum and Outlier) which detects presence of horizontal pleiotropy and accounts for horizontal pleiotropy by correcting for single nucleotide polymorphisms (SNPs) identified as outliers.^(64)^

Detail on the functions used in this analysis can be found in **Supplementary File 1.**

### Multivariable analysis

As previous outlined, MVMR is an extension of MR that allows the estimation of effects of two or more exposures that may be genetically correlated, on an outcome.^(36,37)^ It is advantageous as it enables adjustment for possible pleiotropy that may occur if the exposures have a shared genetic component,^(36,37)^ which may violate the exclusion restriction assumption of UVMR. In addition, MVMR allows adjustment for factors that may be confounders of the association between the genetic variants being used as instruments for the exposure of interest and the outcome, which would violate the independence assumption. Finally, if the exposures are on the same causal pathway, MVMR can be used to estimate a direct effect independently of a potential mediator.^(65)^

We performed MVMR to evaluate the direct effect, using the IVW method, of ever parous status, age at first birth, age at last birth and number of births on overall, ER positive and ER negative breast cancer risk. We additionally performed MVMR of reproductive factors, that appeared to have an effect on risk of ER negative breast cancer, on breast cancer subtypes (HER2 enriched and triple negative). We used the “ivw_mvmr()” from the MVMR R package.^(66)^ GWAS estimates for age at first birth, age at last birth and number of births were standardized based on the standard deviation of the phenotypic exposure (mean = 0 and SD = 1) prior to performing MVMR.

Factors that were considered for MVMR adjustment were 1) traits we identified have an causal effect on the exposure and therefore could violate the independence assumption 2) traits that we have shown have a genetic correlation with the exposure but for which there is little evidence for a causal relationship with the exposure 3) traits that we have identified as potential mediators.^(32,33)^ We adjusted for reproductive factors, age at menarche and menopause, and finally adiposity measures, we adjusted for each trait in turn. (**Figure 1**)

**Figure 1.**
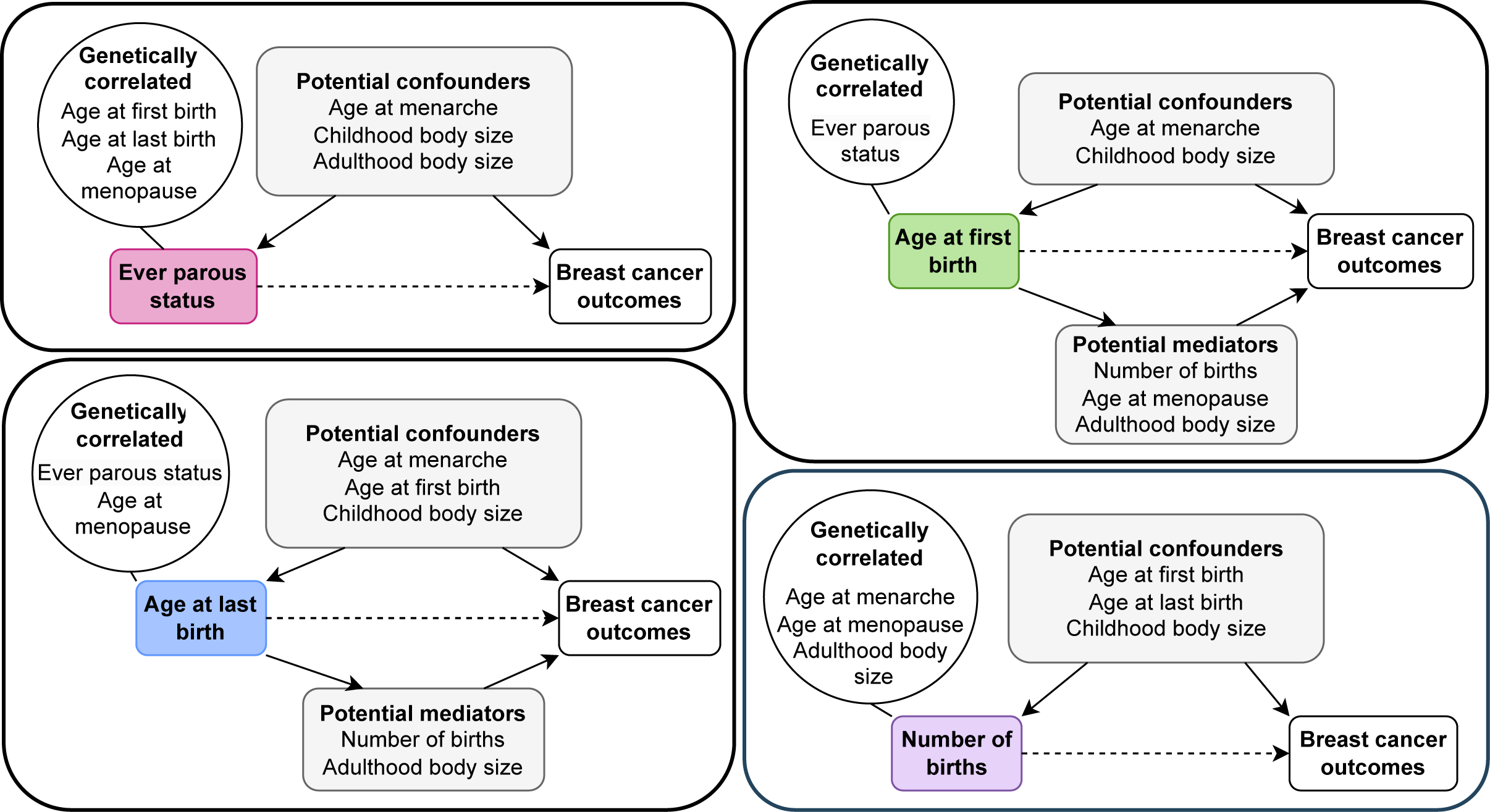
Variables included in multivariable mendelian randomization analysis.

While it may seem implausible to adjust for ever parous status in the MVMR analysis of age at first and last birth, since the age at first and last birth GWAS are conditioned on ever parous status by being performed only on parous women, while the breast cancer risk GWAS is not, it is important to adjust for ever parous status to reduce bias and allow estimation of direct effects.^(67)^

#### Evaluating multivariable mendelian randomization assumptions

We evaluated the instrument strength for the two exposures in the MVMR setting using a conditional F-statistic,^(68)^ and used a modified form of Cochran’s Q statistic to evaluate evidence of horizontal pleiotropy.^(66)^ Where we identify weak instruments and/or evidence of pleiotropy we additionally performed MVMR estimation using Q-statistic minimisation allowing for weak instruments and balanced heterogeneity.^(66)^ We did not use this method when the F statistic falls below 4, since the method does not perform well in this circumstance.^(66)^ We additionally performed the MVMR analysis using the MR Egger method to evaluate the presence of directional pleiotropy.^(69,70)^ This analysis was performed similarly to the initial MVMR analysis in relation to overall, ER positive and ER negative breast cancer risk.

Further details on the functions used to perform these analyses can be found in **Supplementary File 1.**^(66)^

## Results

### UK Biobank

273,238 women from UK Biobank were included. The mean age at assessment was 56 years (SD=8), further sample characteristics are shown in **Table 1**.

**Table 1.**
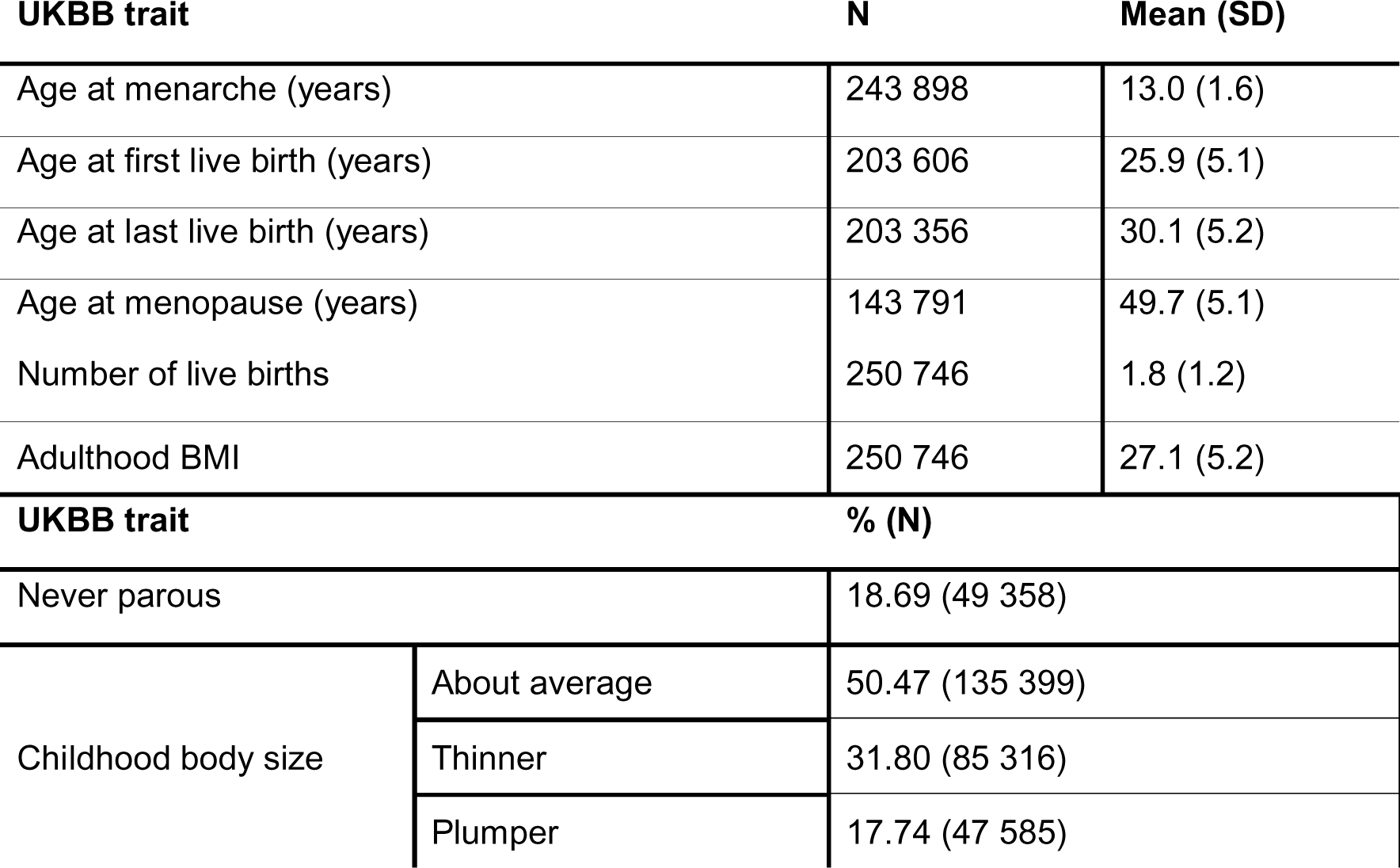
UK Biobank study characteristics included in this study. N=Sample size, SD= Standard deviation. IQR = Interquartile range. UKBB: UK Biobank

#### Genome wide association studies

**Table 2** displays the number of SNPs associated with each reproductive factor at genome-wide significance (p value<5×10^-8)^ after LD clumping and harmonising with the outcome.

**Table 2.** Univariable instrument strength of each trait of interest. N: sample size, nSNPs: number of SNPs.

| Trait | Trait N | Breast cancer outcome | nSNPs | F statistic |
| --- | --- | --- | --- | --- |
| Age at first live birth | 203 606 | Overall | 34 | 39.18 |
|  |  | ER status | 41 | 38.28 |
|  |  | Subtypes | 34 | 39.18 |
| Age at last live birth | 203 356 | Overall | 8 | 34.11 |
|  |  | ER status | 9 | 33.98 |
|  |  | Subtypes | 8 | 34.11 |
| Number of live births | 250 746 | Overall | 8 | 43.64 |
|  |  | ER status | 9 | 42.79 |
|  |  | Subtypes | 8 | 43.64 |
| Ever parous status | 250 746 | Overall | 4 | 39.43 |
|  |  | ER status | 4 | 39.43 |
|  |  | Subtypes | 4 | 39.43 |

#### Genetic correlation

Many of the reproductive factors, adiposity measures, as well as age at menarche and menopause were strongly genetically correlated. Exceptions for the four reproductive factors were: childhood body size and age at first birth, age at last birth and number of births. (**Figure 2, Supplementary File 2: Table S1**) In addition, age at first and last birth were inversely genetically correlated with overall and ER negative breast cancer risk but not ER positive. Ever parous status and number of births were not genetically correlated with any of the breast cancer risk outcomes. (**Figure 2, Supplementary File 2: Table S1**)

**Figure 2.**
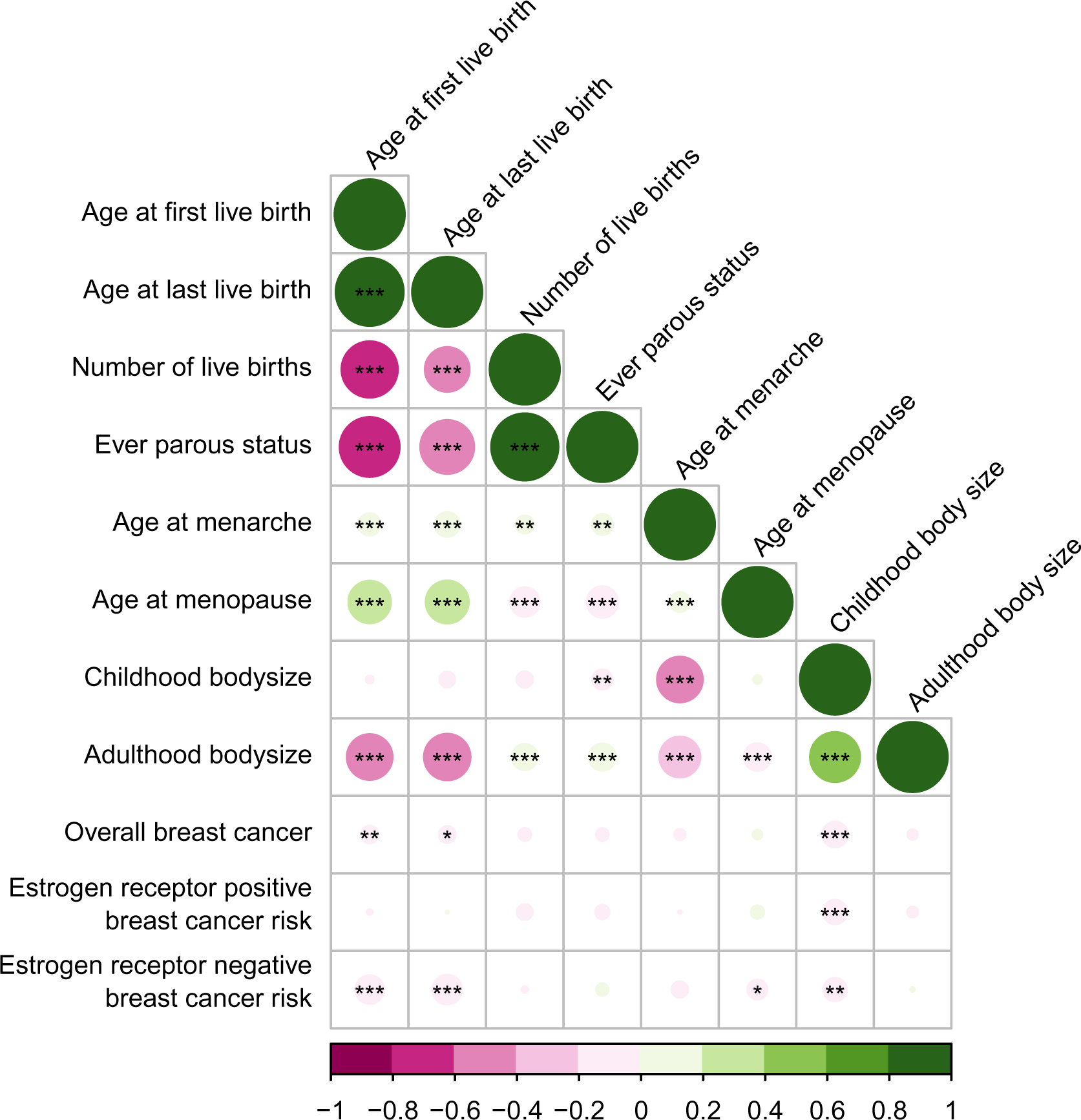
Genetic correlation between reproductive factors, age at menarche and age at menopause, and adiposity measures, and breast cancer risk outcomes. * p-value<0.05, ** p-value<0.01, *** p-value<0.001.

### Estimated causal effects between reproductive factors and adiposity measures

Previous research has shown interrelationships between the four reproductive factors of interest, as well as causal effects between the reproductive factors, adiposity in childhood and adulthood, and age at menarche and menopause (**Figure 3**). Further details on these findings are provided elsewhere.^(32,33)^

**Figure 3.**
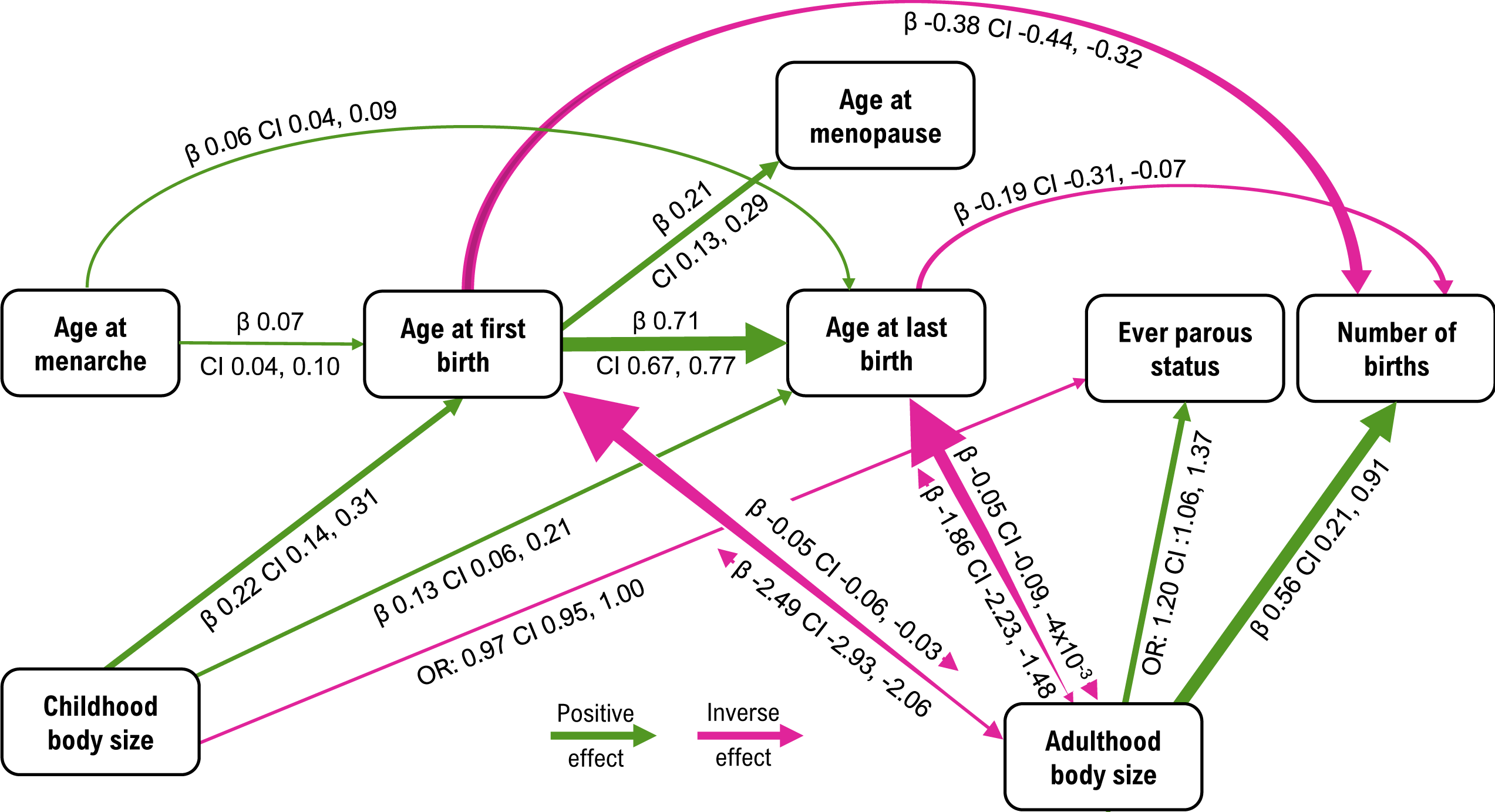
Causal relationships estimated using mendelian randomization in previous work (*Prince et al. 2022, Prince et al. 2023*). OR: Odds ratio, CI: 95% confidence interval. The width of the line/arrowhead indicates the magnitude of the estimate.

### Mendelian randomization

In the univariable MR analysis, all traits had an F statistic over the standard threshold of 10 (**Table 2**). However, in the multivariable analysis the conditional F statistic was reduced for all traits and was below 10 for all reproductive factors investigated; age at first birth, age at last birth, number of births and ever parous status, with adjustment for at least one factors in the MVMR analysis (**Supplementary File 2: Table S1**). We therefore present MVMR estimated using Q-statistic minimisation, which is robust to weak instruments and balanced heterogeneity, as the primary analysis. However, this was not performed where the instruments were deemed too weak, with an F statistic of less than 4. This was the case for age at last birth adjusted for age at menarche, age at menopause, and adulthood body size (each separately); number of births adjusted for age at menarche; and, ever parous status adjusted for age at menarche, age at menopause, age at first birth, childhood and adulthood body size (each separately) (**Supplementary File 2: Table S2**). In these cases, MVMR using the IVW method are presented but these results should be interpreted with caution due to the risk of weak instrument bias. In the MVMR analysis, we adjusted for each factor in turn, thus there were only two exposures: the exposure of interest and the adjustment, and the outcome in each MVMR model.

ORs for age at first birth, age at last birth and number of births are shown as per SD in the phenotypic exposure.

#### Ever parous status

Ever parous status is a binary exposure and therefore, to facilitate interpretation, the effects in **Supplementary File 2: Table S3** and **Figure 4A** have been converted from “ORs per log odds” in the main text here to per doubling in the genetic liability to ever being parous.

**Figure 4.**
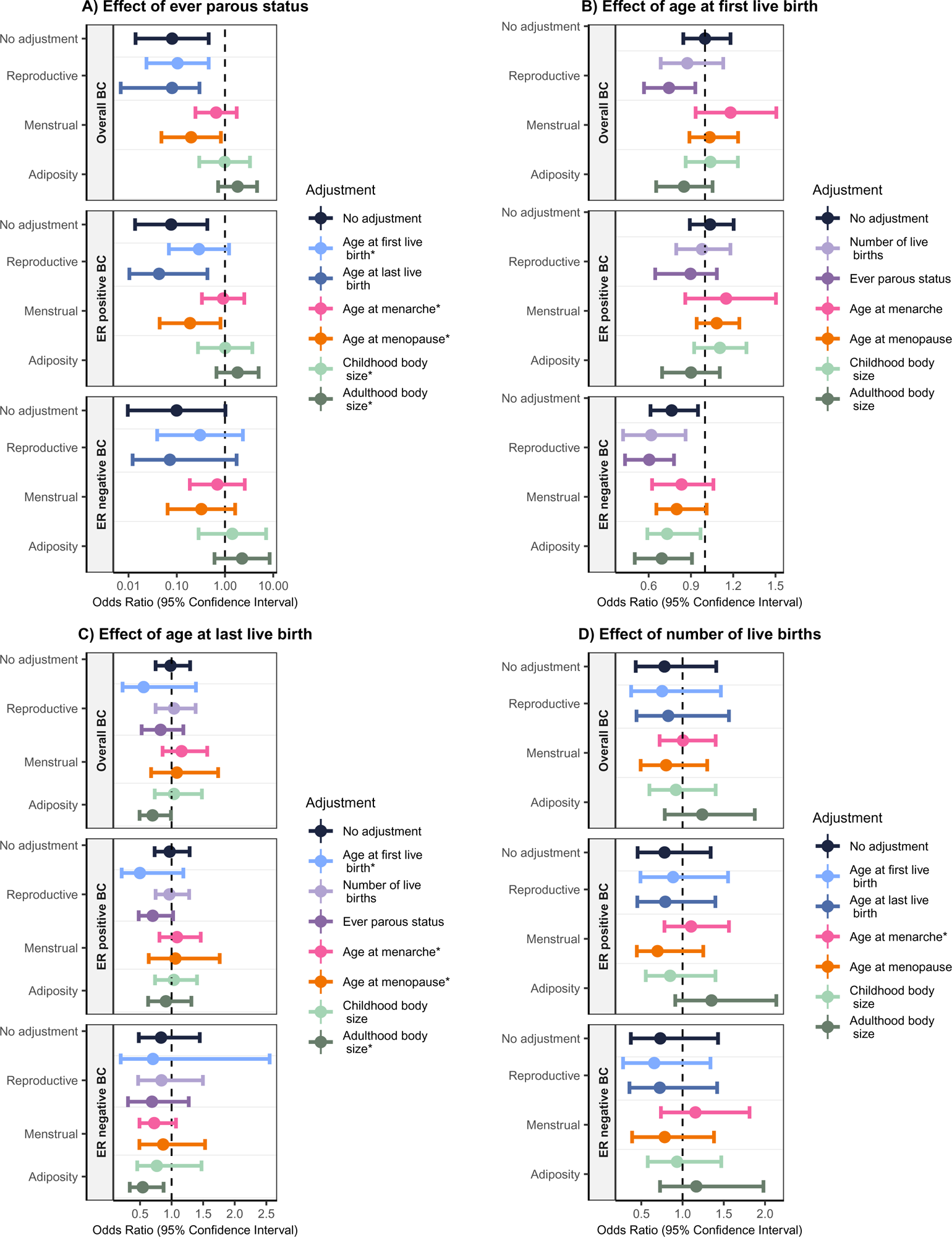
Univariable and multivariable mendelian randomization assessing the effects of A) ever parous status, B) age at first birth, C) age at last birth and D) number of births on overall, ER positive and ER negative breast cancer risk. No adjustment indicated findings from the univariable analysis. Effect of ever parous status is presented per log odds of ever parous status and effects of age at first birth, age at last birth and number of births are presented per standard deviation increase in the phenotypic exposure. * initial MVMR result are presented due to weak instruments preventing the use of MVMR estimated using Q-statistic minimisation.

In the univariable analysis we identified a protective effect of being parous on overall (UVMR: 0.17, CI: 0.05, 0.58), ER positive breast cancer risk (UVMR OR: 0.17, CI: 0.05, 0.56) and ER negative breast cancer risk (UVMR OR: 0.20, CI: 0.04, 1.02).

Multivariable adjustments revealed wide confidence intervals spanning the null for the effect of ever parous status on overall breast cancer risk when adjusted for age at menarche (MVMR OR: 0.74, CI: 0.38, 1.47), childhood body size (MVMR OR: 0.99, CI: 0.43, 2.28) and adulthood body size (MVMR OR: 1.52, CI: 0.80, 2.89); ER positive when adjusted for age at first birth (MVMR OR: 0.42, CI: 0.16, 1.14), age at menarche (MVMR OR: 0.94, CI: 0.47, 1.89), childhood body size (MVMR OR: 1.01, CI: 0.41, 2.47) and adulthood body size (MVMR OR: 1.52, CI: 0.75, 3.05); and ER negative breast cancer risk when adjusted for age at first birth (MVMR OR: 0.44, CI: 0.11, 1.81), age at last birth (MVMR estimated using Q-statistic minimisation OR: 0.16, CI: 0.02, 1.47), age at menarche (MVMR OR: 0.78, CI: 0.31, 1.92), age at menopause (MVMR OR: 0.46, CI: 0.15, 1.40), childhood body size (MVMR OR: 1.27, CI: 0.42, 3.89) and adulthood body size (MVMR OR: 1.76, CI: 0.71, 4.38) (**Figure 4A, Supplementary File 2: Table S3**). It is worth noting that conditional F statistics for ever parous status fell below 4 for all adjustment except age at last birth, and the number of SNPs used as instruments were 4 or less (**Supplementary File 2: Table S2**).

#### Age at first birth

In the univariable analysis we found little evidence for an effect of age at first birth on overall (UVMR OR: 1.00, 95% CI 0.85, 1.18) or ER positive breast cancer risk (UVMR OR: 1.04, CI: 0.89, 1.20) but an inverse effect on ER negative breast cancer risk (UVMR OR: 0.76, CI: 0.61, 0.95). (**Figure 4B, Supplementary File 2: Table S4**)

We identified a protective effect of later age at first birth on overall breast cancer risk after adjusting for ever parous status in the multivariable analysis (MVMR estimated using Q-statistic minimisation OR: 0.74, CI: 0.57, 0.93). Conversely, similarly to the univariable analysis we find minimal evidence of an effect of age at first birth on ER positive breast cancer risk. Although it’s worth noting that adjusting for age at menarche reveals the largest positive effect although confidence intervals are wide, overlapping the univariable estimate and the null (MVMR estimated using Q-statistic minimisation OR: 1.15, CI: 0.86, 1.50). The inverse effect on ER negative breast cancer risk we identified in the univariable analysis somewhat attenuated with adjustment for age at menarche with confidence intervals just crossing the null (MVMR estimated using Q-statistic minimisation OR: 0.83, CI: 0.62, 1.06) and age at menopause (MVMR estimated using Q-statistic minimisation OR: 0.80, CI: 0.66, 1.01). (**Figure 4B, Supplementary File 2: Table S4**)

#### Age at last birth

We found little evidence for an effect of age at last birth on overall (UVMR OR: 0.98, CI: 0.75, 1.29), ER positive (UVMR OR: 0.97, CI: 0.73, 1.29) and ER negative (UVMR OR: 0.83, CI: 0.48, 1.45) breast cancer risk in the univariable analysis (**Figure 4C, Supplementary File 2: Table S5).**

However, we found evidence of an inverse effect of age at last birth adjusting for adulthood body size on overall (MVMR OR:0.70, CI: 0.49, 0.99), and ER negative breast cancer risk (MVMR OR: 0.54, CI: 0.34, 0.87) (**Figure 4C, Supplementary File 2: Table S5).**

Adjusting for age at first birth revealed an inverse effect of larger magnitude on overall (MVMR OR 0.56, CI: 0.22, 1.38) and ER positive breast cancer risk (MVMR OR 0.50, CI: 0.21, 1.19) compared to the univariable analysis. Adjusting for number of births revealed little evidence for an effect of age at last birth on overall, ER positive and ER negative breast cancer risk, similar to the UVMR analysis (**Figure 4C, Supplementary File 2: Table S5).**

#### Number of births

We found limited evidence for an effect of number of births on overall (UVMR OR: 0.78, CI: 0.43, 1.41), ER positive (UVMR OR: 0.78, CI: 0.46, 1.34) or ER negative breast cancer risk (UVMR OR: 0.73, CI: 0.37, 1.43) in the univariable analysis, with a similar result in the multivariable analyses. (**Figure 4D, Supplementary File 2: Table S6**)

#### Estrogen receptor negative subtypes

Since we found some evidence for an effect of age at first birth on ER negative breast cancer risk with relatively strong instruments, we investigated the effects of age at first birth on HER2 enriched and triple negative breast cancer risk. We used UVMR and MVMR in a similar manner as in relation to overall, ER positive and negative breast cancer risk, detail on evaluating the MR assumptions can be found in **Supplementary File 1.**

The univariable analysis revealed limited evidence of an effect of age at first birth on HER2 enriched breast cancer risk (UVMR OR: 0.78, CI:0.47, 1.30), while adjusting for number of births (MVMR estimated using Q-statistic minimisation OR: 0.28, CI: 0.11, 0.57) and ever parous status (MVMR estimated using Q-statistic minimisation OR: 0.38, CI: 0.17, 0.80) revealed inverse effects (**Supplementary File 2: Table S4, Supplementary File 3: Figure S5**). We found minimal evidence for an effect of age at first birth on triple negative breast cancer risk in the UVMR analysis (UVMR OR: 0.88, CI: 0.63, 1.24) which was similar in the MVMR analysis (**Supplementary File 2: Table S4, Supplementary File 3: Figure S5**).

#### Evaluating univariable mendelian randomization assumptions

There was consistency between the univariable analysis using the IVW method and the additional MR methods (MR Egger, weighted median and weighted mode) for the effects of age at last birth and number of births. However, there were inconsistencies with ever parous status and age at first birth. While the effect estimates for ever parous status on overall and ER negative breast cancer risk were largely similar to the IVW method across the additional methods, confidence intervals for the MR Egger and weighted mode spanned the null, with the MR Egger estimate being largely attenuated (**Supplementary File 2: Table S7**). In addition, the effect of age at first birth on ER negative breast cancer risk was similar when using the weighted median and mode methods but not MR Egger, where the estimate was in the positive direction. However, across all additional methods confidence intervals spanned the null (**Supplementary File 2: Table S7**).

Using the MR-PRESSO method, we identified potential outliers for all relationships assessed, except for the effect of age at last birth on overall and ER positive breast cancer risk, and the effect of age at first birth on HER2 enriched and Triple negative breast cancer risk. Correcting for identified outliers didn’t appear to change the evidence for effects identified in the initial analysis using the IVW method. However, it is worth noting that of the 4 SNPs used as instruments for ever parous status, 2 were identified as outliers in relation to overall and ER positive breast cancer risk and 1 in relation to ER negative, meaning this analysis was based on a limited number of instruments (**Supplementary File 2: Table S8**) The Egger intercept test didn’t identified evidence for pleiotropy across any relationships investigated. (**Supplementary File 2: Table S9**)

#### Evaluating multivariable mendelian randomization assumptions

In the multivariable analysis we identified evidence of heterogeneity for all models except for those that included: age at first birth and age at last birth; and, age at last birth and ever parous status in relation to ER positive breast cancer risk, those that included age at first birth and age at last birth in relation to ER negative breast cancer risk, and in the analysis of age at first birth for all adjustments except childhood body size on HER2 enriched breast cancer risk. (**Supplementary File 2: Table S10**). Where there was evidence for heterogeneity and the F statistic was 4 or higher, we performed MVMR estimation using Q-statistic minimisation which is robust to weak instruments and balanced heterogeneity (**Supplementary File 2: Table S3-6**). As previously stated, these results were presented as the primary analysis.

##### MR Egger

We performed the multivariable analysis using the MR Egger method to assess for pleiotropy and the genetic associations with the first exposure were set to be positive. The estimates were mostly consistent with the main analysis, although in many cases, the confidence intervals spanned the null. However, there were inconsistencies. Of note, we identified some evidence for an effect of ever parous status on ER positive breast cancer risk adjusted for age at first birth (OR: 0.30, CI: 0.09, 0.97) which we do not find strong evidence for in the main analysis. (**Supplementary File 2: Table S11, Supplementary File 3: Figure S1**) In addition, the main analysis identified strong evidence for an effect of age at first birth on overall breast cancer risk adjusted for ever parous status which was not seen using the MR Egger method. (**Supplementary File 2: Table S11, Supplementary File 3: Figure S2**) The MR Egger method in the multivariable analysis revealed an inverse effect of number of births, adjusted for age at first birth, on overall (OR: 0.49, CI: 0.26, 0.91) and ER negative breast cancer risk (OR: 0.42, CI: 0.19, 0.91) which was not identified in the main analysis. (**Supplementary File 2: Table S11, Supplementary File 3: Figure S4**)

Where we identified an effect using the IVW method of age at first birth on HER2 enriched breast cancer risk in the multivariable analysis, wide confidence intervals included the null using the MR Egger method (adjusted for ever parous status: OR: 1.08, CI: 0.07, 15.71, adjusted for number of births: OR: 0.27, CI: 0.05, 1.62). (**Supplementary File 2: Table S11**) Although we did identify some evidence for an inverse effect of age at first birth on HER2 enriched breast cancer risk adjusting for age at menopause (OR: 0.46, CI: 0.21, 1.00). (**Supplementary File 2: Table S11**) We additionally identified minimal evidence for an effect of age at first birth on triple negative breast cancer risk in the MVMR analysis using the MR Egger method, similarly to the IVW method. (**Supplementary File 2: Table S11**)

## Discussion

In this study we used univariable, and multivariable Mendelian randomization methods to investigate the role of parity-related reproductive factors on breast cancer development.

Using these approaches, while we found evidence to support that having children has a protective effect on breast cancer risk in the univariable analysis, we could not determine the direct effect independently of age at menarche, age at menopause, reproductive factors and adiposity measures due to the analysis of ever parous status being subject to weak instruments across all the multivariable models. It may be that a larger GWAS of ever parous status is required to use MR more effectively, resulting in a higher number of SNPs identified as robustly associated with ever parous status.

In addition, we found little evidence to support a protective effect of an earlier age at first birth on breast cancer risk.^(2,4–8)^ This may be due to biases such as residual pleiotropy that have driven our MR estimates towards the null. We discuss this limitation in more detail below. However, we do find evidence for an adverse effect of earlier age at first birth on ER negative breast cancer risk in the univariable analysis, although this attenuated somewhat with adjustment for age at menarche and menopause, with confidence intervals just crossing the null. We further evaluated the effect of age at first birth on ER negative breast cancer risk by looking at the HER2 enriched and triple negative subtypes and we find little evidence of an effect in the univariable analysis. However, after adjustment for number of births and ever parous status, we found than earlier age at first birth increases HER2 enriched breast cancer risk.

Since ER negative breast cancer is typically seen in younger, premenopausal women, the increased risk immediately after an earlier first pregnancy is more likely to relate to ER negative breast cancer. This risk is likely to be due to the proliferative effects of pregnancy due to high levels of ovarian hormones which up-regulate proliferation related genes, increasing the risk of mutation.^(9,13)^ Since the effect of age at first birth on breast cancer risk we find is isolated to HER2 enriched breast cancer, it could be that an increased risk of HER2 mutations during pregnancy leads to an overexpression of HER2, driving the development of breast cancer.^(71)^ Additionally, pregnancy is thought to increase breast cancer risk in the shorter-term because of the inflammatory and wound healing properties of mammary involution which occurs after lactation.^(13,15,16)^

Previous observational research has identified that having an age at first birth at or younger than 25 is associated with a lower level of estrogen post menopause compared to nulliparous women.^(72)^ Therefore, it would be valuable for future studies to evaluate whether levels of ovarian hormones such as estrogen and progesterone play a role in the relationship between birth-related factors and breast cancer risk, for example, in a mediation MVMR framework.

Point estimates for the effect of age at last birth across the univariable and multivariable models suggest an adverse effect of a younger age at last birth on ER negative breast cancer risk, which is consistent with a previous MR study,^(31)^ but conflicts with the observational literature.^(7,8,24–26)^

It may seem implausible to adjust for ever parous status in the analysis of age at first and last birth on breast cancer risk outcomes since these events only occur in women who have given birth and therefore ever being parous could not observationally confound or mediate this relationship. However, a recent study has shown that performing MR without adjusting for a factor that was conditioned on in one of the exposure or outcome GWAS can bias the resulting MR estimate, and adjusting for that factor can allow estimation of direct effects.^(67)^ While age at first and last birth are conditioned on ever parous status since the GWAS was for these traits only being performed among parous women, the GWAS of breast cancer risk was not performed solely on parous women or adjusted for ever parous status, and so it is important to adjust for ever parous status in an MVMR model. Moreover, in the analysis of age at first birth on overall breast cancer risk, adjusting for ever parous status did modify the estimate to reveal an inverse effect, compared to the limited evidence identified in the univariable analyses. It would have been informative to perform the analysis of age at first and last birth using GWAS from the BCAC consortium performed solely in parous women, however this data, to our knowledge, is not publicly available.

We find limited evidence that number of births has an impact on breast cancer risk in either the univariable or multivariable analysis, which is consistent with a previous MR study.^(30)^

Due to weak instruments, we cannot make any definitive conclusions regarding age at last birth or ever parous status for most MVMR models. Where instruments were at least slightly stronger, with a F statistic of 4 or higher, in the multivariable analyses, we performed MVMR estimation using Q-statistic minimisation, and the direction and strength of evidence was largely similar to the UVMR model. While weak instruments prevent in depth speculation, in the case of age at last birth, it may be that we are capturing the effect of age at first birth since these factors are highly genetically correlated. Indeed, in the multivariable analysis of age at last birth on breast cancer risk outcomes adjusted for age at first birth confidence intervals were particularly wide, compared to the other models with age at last birth as the exposure.

We used the MR Egger method to assess for evidence of pleiotropy, for the most part, this method revealed similar evidence to the main analysis. However, we did not identify evidence for the inverse effect of age at first birth on overall breast cancer risk, adjusting for ever parous status in the MVMR analysis using the MR Egger method which we did find in the main analysis. In addition, using the MR Egger method, adjusting for age at first birth uncovered evidence that a higher number of children leads to reduced overall and ER negative breast cancer risk. While this suggests the main analysis may be biased by pleiotropy, the instrument strength was deemed weak in the multivariable analyses, with an F statistic below 10, which limited confidence in these results and prevented further assessment of pleiotropy using robust methods.^(73)^

### Strengths

The main strength of this study is the use of a multivariable MR approach allowing the evaluation of direct effects of reproductive factors on breast cancer risk outcomes. There has not previously been an informed, systematic MVMR study, using information from previous work on genetic correlation and causal relationships between the reproductive factors of interest and age at menarche and menopause, adiposity measures and other reproductive factors.^(32,33)^

Additionally, this study uses the UK Biobank study which has phenotypic and genetic data on a large number of individuals which increases the likelihood of identifying strong genetic instruments for each exposure of interest. Another strength is the use of GWAS data from the BCAC consortium, allowing the investigation of risk of breast cancer by ER status, and risk of breast cancer subtypes.

### Limitations

This study has a number of limitations that should be considered when interpreting results. Firstly, weak instruments were a key limitation when investigating the effects of age at last birth and ever parous status on breast cancer risk outcomes, especially in the multivariable models. This can be attributed to the genetic correlation that is present between these traits and the other factors included in the multivariable analyses,^(32)^ as well as the low number of SNPs that arise as genome-wide significant in relation to these traits (8 and 4 for age at last birth and ever parous status, respectively). Where the instrument strength was not deemed too weak, with an F statistic of 4 or higher, we additionally performed MVMR estimation using Q-statistic minimisation that is developed to be robust to weak instruments. However, this was not possible for number of births adjusted for age at menarche, and all MVMR models for ever parous status, except with adjustment for age at last birth, and additionally for age at last birth, except with adjustment for number of births, ever parous status and childhood body size. Furthermore, it would have been useful to include multiple adjustments within the MVMR models, however instruments were too weak to facilitate this. Larger GWAS and stronger instruments are required to extend our MVMR approach, however it may be that, due to the aforementioned genetic correlation, an exceptionally large sample size would be required.

Secondly, as previously mentioned, we may not find a protective effect of an early age at first birth on breast cancer risk because of biases due to violations of the MR assumptions. It is plausible that we may be capturing residual pleiotropic effects as a result of age at menarche confounding the relationship between age at first birth and breast cancer risk. The univariable analysis reveals little evidence for an effect of age at first birth on ER positive breast cancer risk (OR: 1.04, CI: 0.89, 1.20 per SD increase in age at first birth), while the effect in the multivariable analysis, adjusted for age at menarche, is larger in the protective direction, although confidence intervals crossed the null (OR: 1.18, CI: 0.93, 1.51 per SD increase in age at first birth). Given the conditional F statistics in this MVMR model (age at first birth: 5.3, age at menarche: 25.0) and that age at menarche and first birth are strongly genetically correlated, the effect in the multivariable analysis may be biased by weak instruments or from pleiotropy via another trait. Stronger instruments for age at first birth in an MVMR model adjusted for age at menarche might reveal the protective effect seen in the observational literature.

Thirdly, data on the reproductive factors investigated have been derived from self-report which can be unreliable. However, we can be relatively confident that reports of age at first and last birth and number of births are accurate since these are significant life events that are likely to be reliably recalled. Additionally, there has typically been good replication of the genetic scores for the reproductive traits in other cohorts.^(74–76)^

In addition, since the reproductive factors investigated in this study are bio-social, findings may not be generalisable to other non-UK populations that have different social norms. Additionally, the findings may not be generalisable to younger populations. It would have been valuable to compare the findings in the study, where the population was predominately European, to populations with other ancestries such as Asian and African. However, while Asian and African ancestry GWAS summary statistics for breast cancer risk are available, those for the four reproductive factors investigated here are not, to the best of our knowledge.

Finally, the MR analysis performed assumes a constant effect of age at first and last birth, and number of births, on breast cancer risk. Future studies could use MR to evaluate the effects of an early age at first birth compared to an average and late age, and vice versa. This approach would also enable the evaluation of whether there is a cross-over point where breast cancer risk changes from parous women to nulliparous women at a certain age at first birth as previously proposed.^(14,15)^ Again, further analysis would require the identification of additional and stronger genetic instruments.

## Conclusion

Using a series of univariable and multivariable MR analyses, we found minimal evidence for a protective effect of early age at first birth on breast cancer risk that has been identified in observational studies, which may be due to unmeasured biases. On the other hand, we identify some evidence of an adverse effect on ER negative breast cancer risk, which supports the notion that there is an immediate increase in breast cancer risk post pregnancy. Understanding of mechanism behind ER negative breast cancers is important since it has worse prognosis compared with those that are ER positive.

In addition, we show the number of births that women have has little effect on overall, ER positive or ER negative breast cancer risk. Future studies considering non-linear relationships between age at first birth and breast cancer risk may provide additional insights.

While our findings for ever parous status and age at last birth do not all concur with the existing literature from non-genetic studies, this may partly be due to limitations of the MVMR approach, specifically relating to instrument strength, which may be overcome with larger GWAS of these traits.

## Authors’ Contributions

C.P. was responsible for Analysis, Investigation and Writing - Original Draft. E.S., G.C.S., L.D.H., A.F., B.L.-L. and R.C.R. were responsible for Conceptualization, Writing - Review & Editing and Supervision. R.C.R. and B.L.-L. were additionally responsible for Investigation. All authors read and approved the final manuscript.

## Data Availability

The availability of all data analysed in this study has been referenced throughout the manuscript and supplementary materials.
GWAS summary statistics for breast cancer risk outcomes were obtained from https://bcac.ccge.medschl.cam.ac.uk/bcacdata/

## Acknowledgements

This research has been conducted using the UK Biobank Resource under Application Number 6326. We thank the participants and researchers from the UK Biobank who contributed or collected data.

The breast cancer genome-wide association analyses for BCAC and CIMBA were supported by Cancer Research UK (PPRPGM-Nov20\100002, C1287/A10118, C1287/A16563, C1287/A10710, C12292/A20861, C12292/A11174, C1281/A12014, C5047/A8384, C5047/A15007, C5047/A10692, C8197/A16565) and the Gray Foundation, The National Institutes of Health (CA128978, X01HG007492-the DRIVE consortium), the PERSPECTIVE project supported by the Government of Canada through Genome Canada and the Canadian Institutes of Health Research (grant GPH-129344) and the Ministère de l’Économie, Science et Innovation du Québec through Genome Québec and the PSRSIIRI-701 grant, the Quebec Breast Cancer Foundation, the European Community’s Seventh Framework Programme under grant agreement n° 223175 (HEALTH-F2-2009-223175) (COGS), the European Union’s Horizon 2020 Research and Innovation Programme (634935 and 633784), the Post-Cancer GWAS initiative (U19 CA148537, CA148065 and CA148112 - the GAME-ON initiative), the Department of Defence (W81XWH-10-1-0341), the Canadian Institutes of Health Research (CIHR) for the CIHR Team in Familial Risks of Breast Cancer (CRN-87521), the Komen Foundation for the Cure, the Breast Cancer Research Foundation and the Ovarian Cancer Research Fund.

## Ethics

UK Biobank received ethical approval from the North West Multi-Centre Research Ethics Committee (REC reference: 16/NW/0274) and was conducted in accordance with the principles of the Declaration of Helsinki.

## Availability of data and materials

The availability of all data analysed in this study has been referenced throughout the manuscript and supplementary materials.

GWAS summary statistics for breast cancer risk outcomes were obtained from https://bcac.ccge.medschl.cam.ac.uk/bcacdata/

