## Supplementary File 1 for "Evaluating the causal impact of reproductive factors on breast cancer risk: a multivariable mendelian randomization approach"

### Methods

#### UK Biobank variables

##### Reproductive factors

To identify the number of live births, women were asked "How many children have you given birth to? (Please include live births only)". A binary measure of parous status, coded as “0” or “1”, where women who had or had not given birth, was derived from the measure of number of live births. Women who indicated that they had given birth to one child were asked "How old were you when you had your child?". Women who indicated that they had given birth to more than one child were asked "How old were you when you had your FIRST child?" and "How old were you when you had your LAST child?". To derive age at first live birth and age at last live birth, responses from primiparous and multiparous women were combined.

##### Menstrual traits

Age at menarche was derived from the question: "How old were you when your periods started?". Age at menopause was derived from the question "How old were you when your periods stopped?", this did not include women who had a hysterectomy/were not sure whether they had gone through menopause.

##### Adiposity measures

A genetic score of the childhood body size measure has been validated as a predictor of childhood adiposity by previous studies in 3 European cohorts; Trøndelag Health Study,^(1)^ The Young Finn Study,^(2)^ and the Avon Longitudinal Study of Parents and Children.^(3)^ In addition, the polygenic score for childhood body size in UK Biobank was more correlated with childhood obesity in an independent sample compared to the adulthood body mass index (BMI) genome-wide association studies (GWAS).^(4)^ Finally genetic risk scores for childhood body size are more strongly associated with fat mass compared to lean mass.^(5)^

#### GWAS

BOLT-LMM was used to conduct the analysis in the GWAS pipeline,^(6)^ which accounts for population stratification and relatedness using linear mixed modelling. Genotyping chip and age were included as covariates. Genome-wide significant single nucleotide polymorphisms (SNPs) were selected at p <5×10^−8^ and were clumped to ensure independence at linkage disequilibrium (LD) r^2^ < 0.001 and a distance of 10 000 kb using the TwoSampleMR package.^(7)^

#### Genetic correlation

Genetic correlation was performed using the linkage disequilibrium score regression (LDSC). The regressions were performed using pre-computed LD scores for each SNP calculated based on individuals of European ancestry from 1000 Genomes European data.^(8)^ These LD scores were filtered to HapMap3 SNPs as these are well-imputed in most studies.^(9)^ SNPs found on chromosome 6 in the region 26MB to 34MB were excluded. GWAS summary statistics were converted for LDSC regression using the munge_sumstats.py command from the command line tool “ldsc”, and LDSC was performed using the ldsc.py command.

#### Univariable mendelian randomization

To evaluate evidence of pleiotropy we applied the function “mr()” to perform MR using the weighted mode, weighted median and MR Egger methods, we additionally applied the “pleiotropy_mr()” function to performed the MR Egger intercept test.^(7,10)^ Furthermore, we applied the “mr_presso” function from the “rondolab/MR-PRESSO” R package.^(11)^

#### Multivariable mendelian randomization

To evaluate the conditional F statistic, horizontal pleiotropy, and MVMR estimation using Q-statistic minimisation we applied the ‘strength_mvmr()’, ‘pleiotropy_mvmr()’, and ‘qhet_mvmr()’ functions from the “MVMR” R package.^(12)^ We calculated the phenotypic correlation between exposures in using the the MVMR models using Pearson’s correlation coefficient which was required for applying the ‘strength_mvmr()’,  ‘pleiotropy_mvmr()’ and ‘qhet_mvmr()’ functions from the “MVMR” R package.^(12)^

Additionally we applied the “mr_mvegger” function from the “MendelianRandomization” R package to evaluate evidence of horizontal pleiotropy, and the genetic associations with the first exposure were set to positive.^(13)^

### Results

#### Estrogen receptor negative subtypes

The F statistic for age at first birth in the univariable analysis was 39.18. However, in the multivariable analysis the F statistic was reduced, falling below 10 with adjustment for at least one of the factors but above 4 allowing use to perform MVMR estimation using Q-statistic minimisation (**Table S2**). There was evidence of heterogeneity in the MVMR analysis of age at first birth on triple negative breast cancer, but limited evidence on HER2 enriched breast cancer (**Table S12**).

*MR Egger*

We performed UVMR and MVMR using the MR Egger method of age at first birth on HER2 enriched and triple negative breast cancer. The MR Egger method revealed a similar effect of age at first birth adjusted for HER2 enriched breast cancer however confidence intervals spanned the null. We identified limited evidence when adjusting for ever parous status, conflicting with the inverse effect found in the main analysis. Additionally, the MR Egger method revealed evidence for an effect of age at first birth on HER2 enriched breast cancer adjusted for age at menopause which was not identified in the main analysis. The MR Egger method revealed limited evidence for an effect of age at first birth on triple negative breast cancer (**Table S14**).

### STROBE-MR checklist of recommended items to address in reports of Mendelian randomization studies^1^ ^2^

| **Item No.** | **Section** | **Checklist item** | **Check** | **Page No.** | **Relevant text from manuscript** |
| --- | --- | --- | --- | --- | --- |
| 1 | **TITLE and ABSTRACT** | Indicate Mendelian randomization (MR) as the study’s design in the title and/or the abstract if that is a main purpose of the study | ☒ | Main text: page 1 | “Evaluating the causal impact of reproductive factors on breast cancer risk: a multivariable mendelian randomization approach” |
|  | **INTRODUCTION** |  |  |  |  |
| 2 | **Background** | Explain the scientific background and rationale for the reported study. What is the exposure? Is a potential causal relationship between exposure and outcome plausible? Justify why MR is a helpful method to address the study question | ☒ | Main text: page 2-5 | “It is currently unclear how each reproductive event affects risk in isolation since these traits are highly correlated with, and/or causally linked to other reproductive factors as well as age at menarche and menopause, and adiposity measures, all established breast cancer risk factors.” |
| 3 | **Objectives** | State specific objectives clearly, including pre-specified causal hypotheses (if any). State that MR is a method that, under specific assumptions, intends to estimate causal effects | ☒ | Main text: page 5 | “We aimed to use univariable and multivariable MR methods to untangle the effects of ever having children, age at first birth, age at last birth and number of children birthed on breast cancer risk. We further aimed to investigate whether effects are independent of age at menarche and menopause, and adiposity measures as well as other reproductive factors, and whether effects differ for ER positive compared to ER negative breast cancer. Furthermore, based on these findings we aimed to assess the effects of reproductive factors on intrinsic breast cancer subtypes.” |
|  | **METHODS** |  |  |  |  |
| 4 | **Study design and data sources** | Present key elements of the study design early in the article. Consider including a table listing sources of data for all phases of the study. For each data source contributing to the analysis, describe the following: |  |  |  |
|  | a) | Setting: Describe the study design and the underlying population, if possible. Describe the setting, locations, and relevant dates, including periods of recruitment, exposure, follow-up, and data collection, when available. | ☒ | Main text: page 6 |  |
|  | b) | Participants: Give the eligibility criteria, and the sources and methods of selection of participants. Report the sample size, and whether any power or sample size calculations were carried out prior to the main analysis | ☒ | Main text: page 6 |  |
|  | c) | Describe measurement, quality control and selection of genetic variants | ☒ | Supplementary File 1: page 1-2 |  |
|  | d) | For each exposure, outcome, and other relevant variables, describe methods of assessment and diagnostic criteria for diseases | ☒ | Supplementary File 1: page 1-2, main text: page 6-8 |  |
|  | e) | Provide details of ethics committee approval and participant informed consent, if relevant | ☒ | Main text: page 26 |  |
| 5 | **Assumptions** | Explicitly state the three core IV assumptions for the main analysis (relevance, independence and exclusion restriction) as well assumptions for any additional or sensitivity analysis | ☒ | Main text: page 9 |  |
| 6 | **Statistical methods: main analysis** | Describe statistical methods and statistics used |  |  |  |
|  | a) | Describe how quantitative variables were handled in the analyses (i.e., scale, units, model) | ☒ | Main text: page 9, 10 | “GWAS estimates were standardized based on the standard deviation of the phenotypic exposure (mean = 0 and standard deviation (SD) = 1) prior to performing MR.” |
|  | b) | Describe how genetic variants were handled in the analyses and, if applicable, how their weights were selected | ☐ | Supplementary File 1: page 2 | “Genome-wide significant single nucleotide polymorphisms (SNPs) were selected at p <5×10−8 and were clumped to ensure independence at linkage disequilibrium (LD) r2 < 0.001 and a distance of 10 000 kb using the TwoSampleMR package.” |
|  | c) | Describe the MR estimator (e.g. two-stage least squares, Wald ratio) and related statistics. Detail the included covariates and, in case of two-sample MR, whether the same covariate set was used for adjustment in the two samples | ☒ | Main text: page 9, 10 | “The primary analysis focused on the inverse variance weighted (IVW) MR method” |
|  | d) | Explain how missing data were addressed | n/a |  |  |
|  | e) | If applicable, indicate how multiple testing was addressed | n/a |  |  |
| 7 | **Assessment of assumptions** | Describe any methods or prior knowledge used to assess the assumptions or justify their validity | ☒ | Main text: page 9-11 |  |
| 8 | **Sensitivity analyses and additional analyses** | Describe any sensitivity analyses or additional analyses performed (e.g. comparison of effect estimates from different approaches, independent replication, bias analytic techniques, validation of instruments, simulations) | n/a |  |  |
| 9 | **Software and pre-registration** |  |  |  |  |
|  | a) | Name statistical software and package(s), including version and settings used | ☒ | Main text: page 9,10, Supplementary File 1: page 2,3 |  |
|  | b) | State whether the study protocol and details were pre-registered (as well as when and where) | n/a |  |  |
|  | **RESULTS** |  |  |  |  |
| 10 | **Descriptive data** |  |  |  |  |
|  | a) | Report the numbers of individuals at each stage of included studies and reasons for exclusion. Consider use of a flow diagram | ☒ | Table 1 |  |
|  | b) | Report summary statistics for phenotypic exposure(s), outcome(s), and other relevant variables (e.g. means, SDs, proportions) | ☒ | Table 2 |  |
|  | c) | If the data sources include meta-analyses of previous studies, provide the assessments of heterogeneity across these studies | n/a |  |  |
|  | d) | For two-sample MR:  i.  Provide justification of the similarity of the genetic variant-exposure associations between the exposure and outcome samples  ii.  Provide information on the number of individuals who overlap between the exposure and outcome studies | ☒  No overlap | Main text: page 6-8 |  |
| 11 | **Main results** |  |  |  |  |
|  | a) | Report the associations between genetic variant and exposure, and between genetic variant and outcome, preferably on an interpretable scale | ☒ | Supplementary table S12-15 |  |
|  | b) | Report MR estimates of the relationship between exposure and outcome, and the measures of uncertainty from the MR analysis, on an interpretable scale, such as odds ratio or relative risk per SD difference | ☒ | Main text: page 13-15,17,18, Figure 4 |  |
|  | c) | If relevant, consider translating estimates of relative risk into absolute risk for a meaningful time period | n/a |  |  |
|  | d) | Consider plots to visualize results (e.g. forest plot, scatterplot of associations between genetic variants and outcome versus between genetic variants and exposure) | ☒ | Figure 4 |  |
| 12 | **Assessment of assumptions** |  |  |  |  |
|  | a) | Report the assessment of the validity of the assumptions | ☒ | Main text: page 14,15, 19-20 |  |
|  | b) | Report any additional statistics (e.g., assessments of heterogeneity across genetic variants, such as *I^2^*, Q statistic or E-value) | ☒ | Supplementary table 10 |  |
| 13 | **Sensitivity analyses and additional analyses** |  |  |  |  |
|  | a) | Report any sensitivity analyses to assess the robustness of the main results to violations of the assumptions | ☒ | Main text: page 15-17 |  |
|  | b) | Report results from other sensitivity analyses or additional analyses | n/a |  |  |
|  | c) | Report any assessment of direction of causal relationship (e.g., bidirectional MR) | n/a |  |  |
|  | d) | When relevant, report and compare with estimates from non-MR analyses | n/a |  |  |
|  | e) | Consider additional plots to visualize results (e.g., leave-one-out analyses) | n/a |  |  |
|  | **DISCUSSION** |  |  |  |  |
| 14 | **Key results** | Summarize key results with reference to study objectives | ☒ | Main text: page 18 |  |
| 15 | **Limitations** | Discuss limitations of the study, taking into account the validity of the IV assumptions, other sources of potential bias, and imprecision. Discuss both direction and magnitude of any potential bias and any efforts to address them | ☒ | Main text: page 21-23 |  |
| 16 | **Interpretation** |  |  |  |  |
|  | a) | Meaning: Give a cautious overall interpretation of results in the context of their limitations and in comparison with other studies | ☒ | Main text: page 21 |  |
|  | b) | Mechanism: Discuss underlying biological mechanisms that could drive a potential causal relationship between the investigated exposure and the outcome, and whether the gene-environment equivalence assumption is reasonable. Use causal language carefully, clarifying that IV estimates may provide causal effects only under certain assumptions | ☒ | Main text: page 19 |  |
|  | c) | Clinical relevance: Discuss whether the results have clinical or public policy relevance, and to what extent they inform effect sizes of possible interventions | n/a |  |  |
| 17 | **Generalizability** | Discuss the generalizability of the study results (a) to other populations, (b) across other exposure periods/timings, and (c) across other levels of exposure | ☒ | Main text: page 23 |  |
|  | **OTHER INFORMATION** |  |  |  |  |
| 18 | **Funding** | Describe sources of funding and the role of funders in the present study and, if applicable, sources of funding for the databases and original study or studies on which the present study is based | ☒ | Main text: page 24-25 |  |
| 19 | **Data and data sharing** | Provide the data used to perform all analyses or report where and how the data can be accessed, and reference these sources in the article. Provide the statistical code needed to reproduce the results in the article, or report whether the code is publicly accessible and if so, where | ☒ | Main text: page 26 |  |
| 20 | **Conflicts of Interest** | All authors should declare all potential conflicts of interest | ☒ | Main text: page 24 |  |

This checklist is copyrighted by the Equator Network under the Creative Commons Attribution 3.0 Unported (CC BY 3.0) license.

1.	Skrivankova VW, Richmond RC, Woolf BAR, Yarmolinsky J, Davies NM, Swanson SA, et al. Strengthening the Reporting of Observational Studies in Epidemiology using Mendelian Randomization (STROBE-MR) Statement. JAMA. 2021;under review.

2.	Skrivankova VW, Richmond RC, Woolf BAR, Davies NM, Swanson SA, VanderWeele TJ, et al. Strengthening the Reporting of Observational Studies in Epidemiology using Mendelian Randomisation (STROBE-MR): Explanation and Elaboration. BMJ. 2021;375:n2233.
