## Supplementary File 3 for "Evaluating the causal impact of reproductive factors on breast cancer risk: a multivariable mendelian randomization approach"

Figure S1 **Multivariable mendelian randomization using MR Egger assessing the effects of ever parous status on overall, ER positive and ER negative breast cancer risk.**

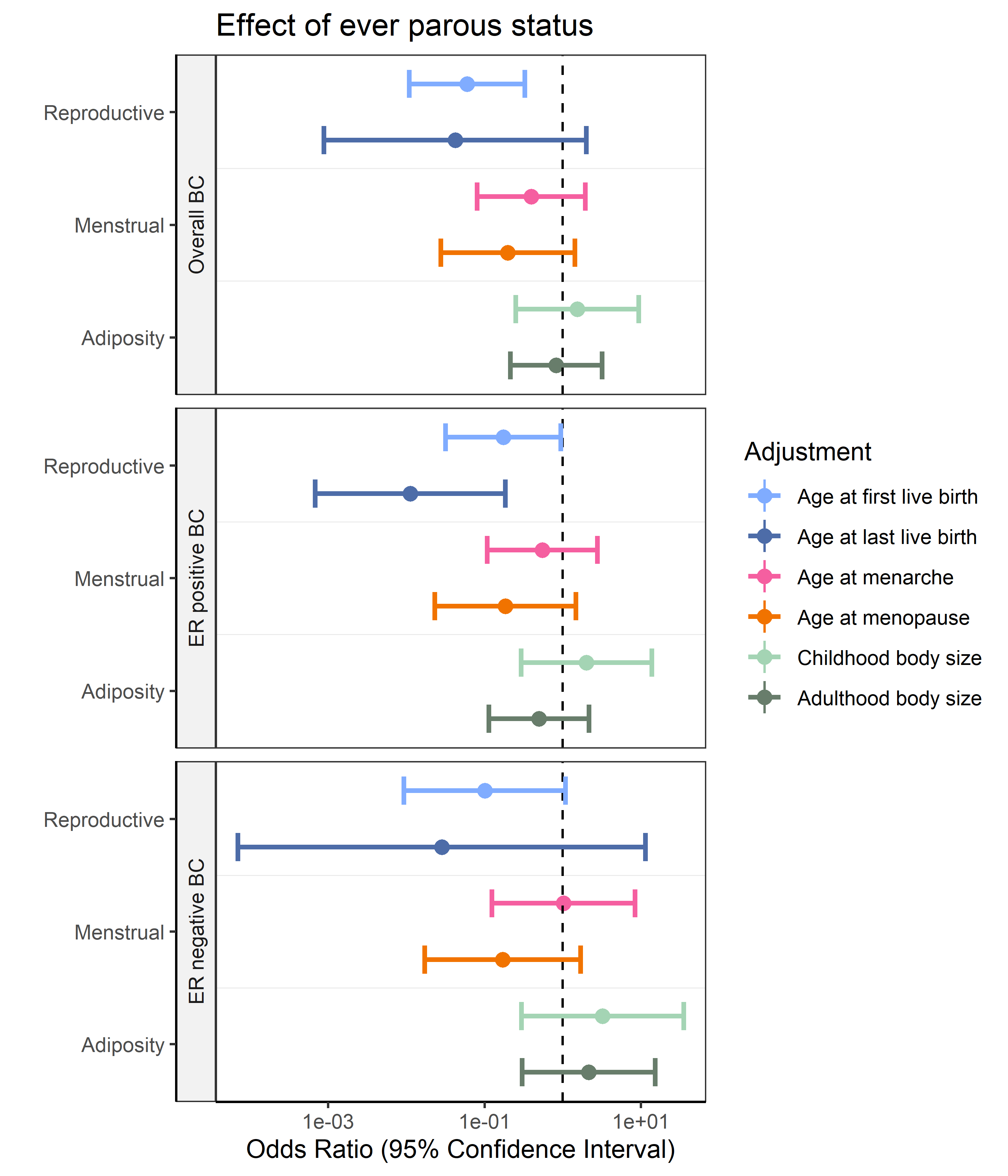

Figure S2 **Multivariable mendelian randomization using MR Egger assessing the effects of age at first birth on overall, ER positive and ER negative breast cancer risk.**

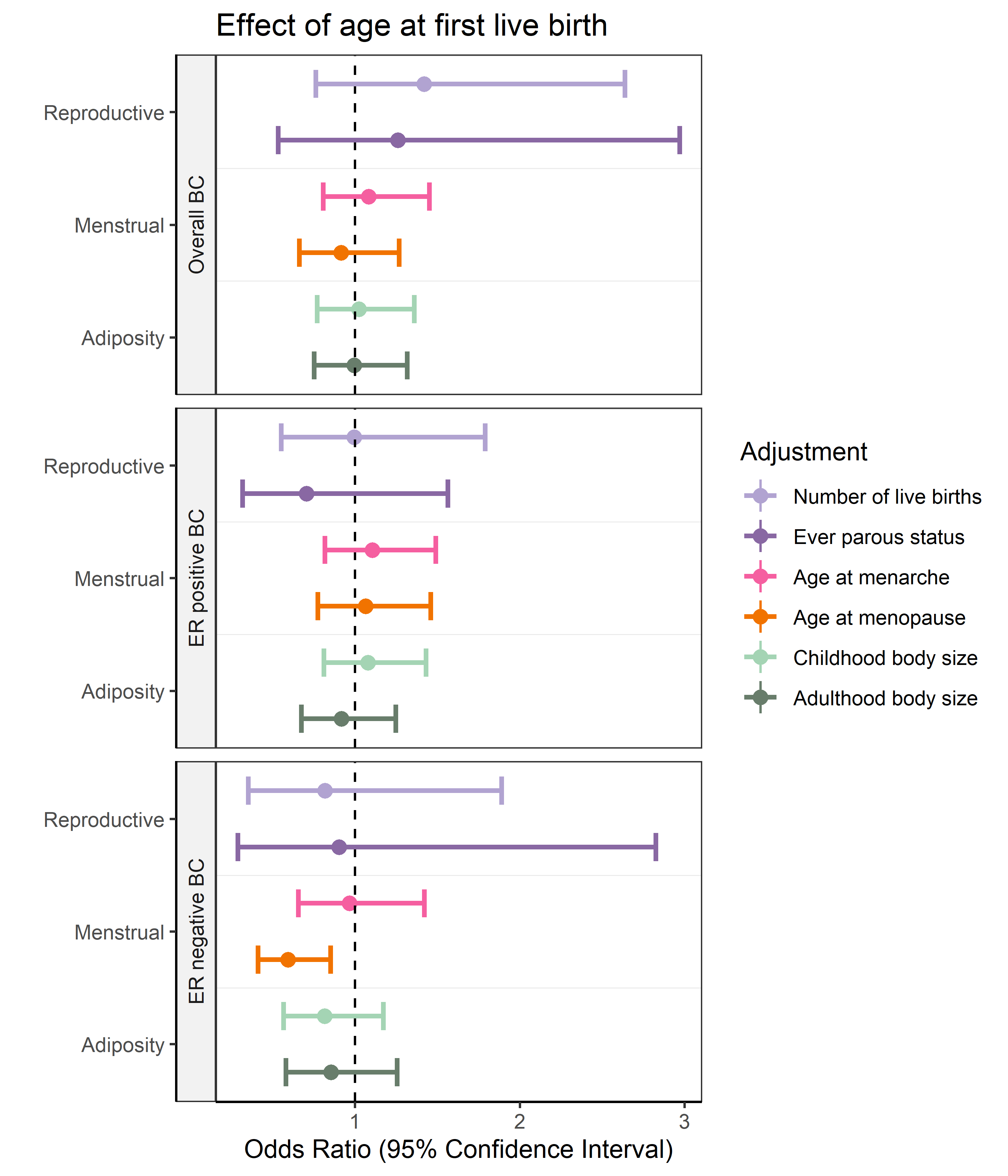

Figure S3 **Multivariable mendelian randomization using MR Egger assessing the effects of age at last live births on overall, ER positive and ER negative breast cancer risk**.

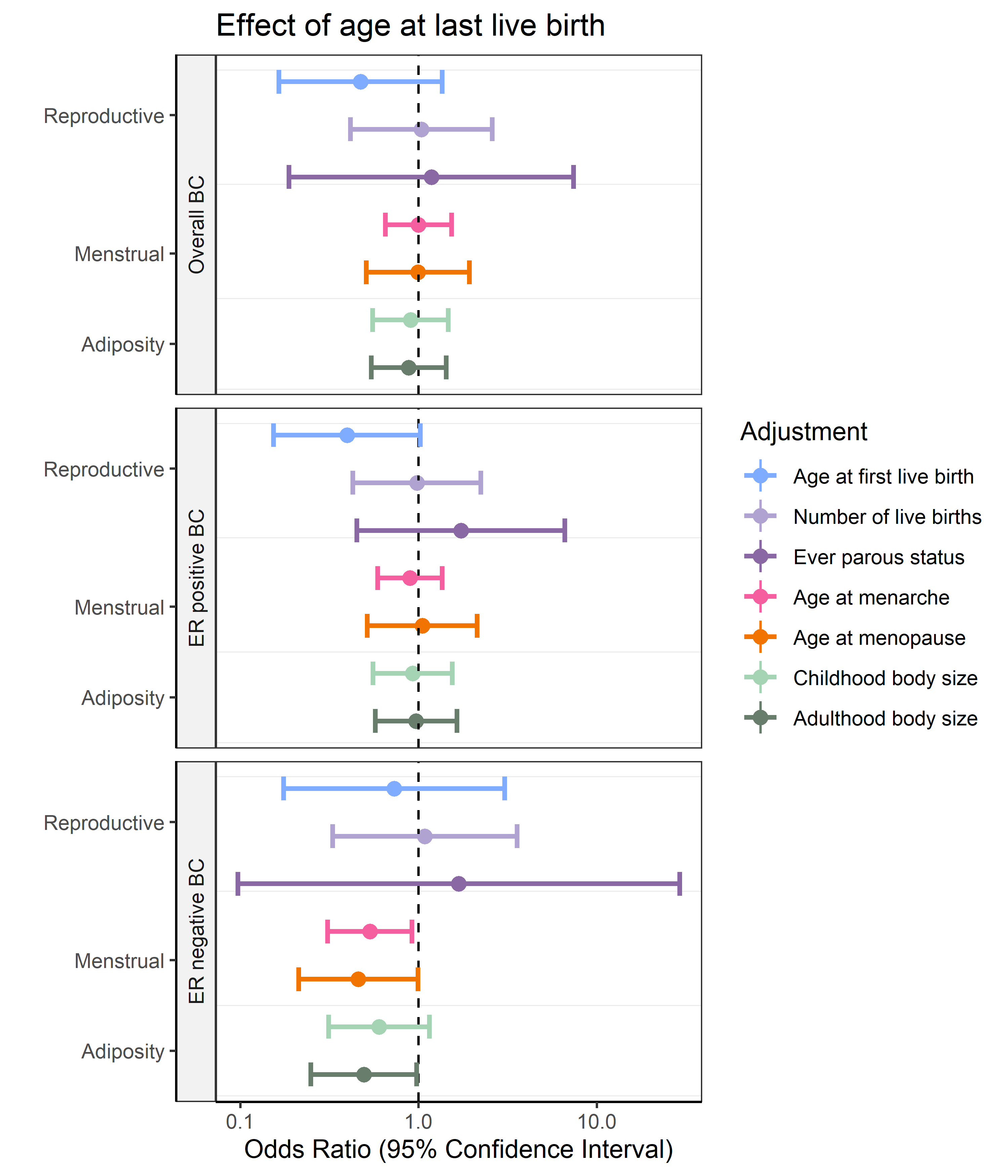

Figure S4 **Multivariable mendelian randomization using MR Egger assessing the effects of number of births on overall, ER positive and ER negative breast cancer risk.**

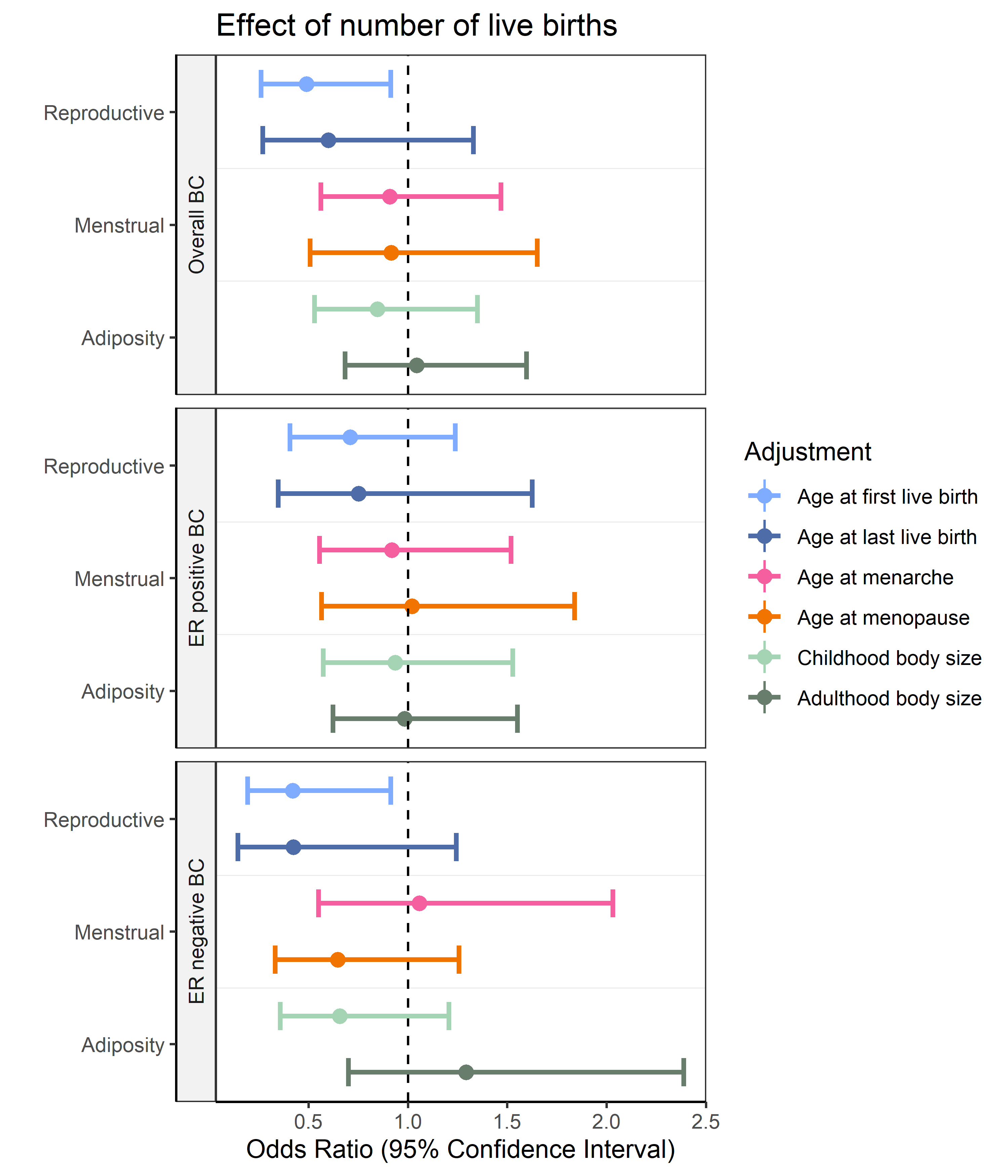

Figure S5 **Univariable and multivariable mendelian randomization assessing the effects of age at first birth, on HER2 enriched and triple negative breast cancer risk.** No adjustment indicated findings from the univariable analysis.

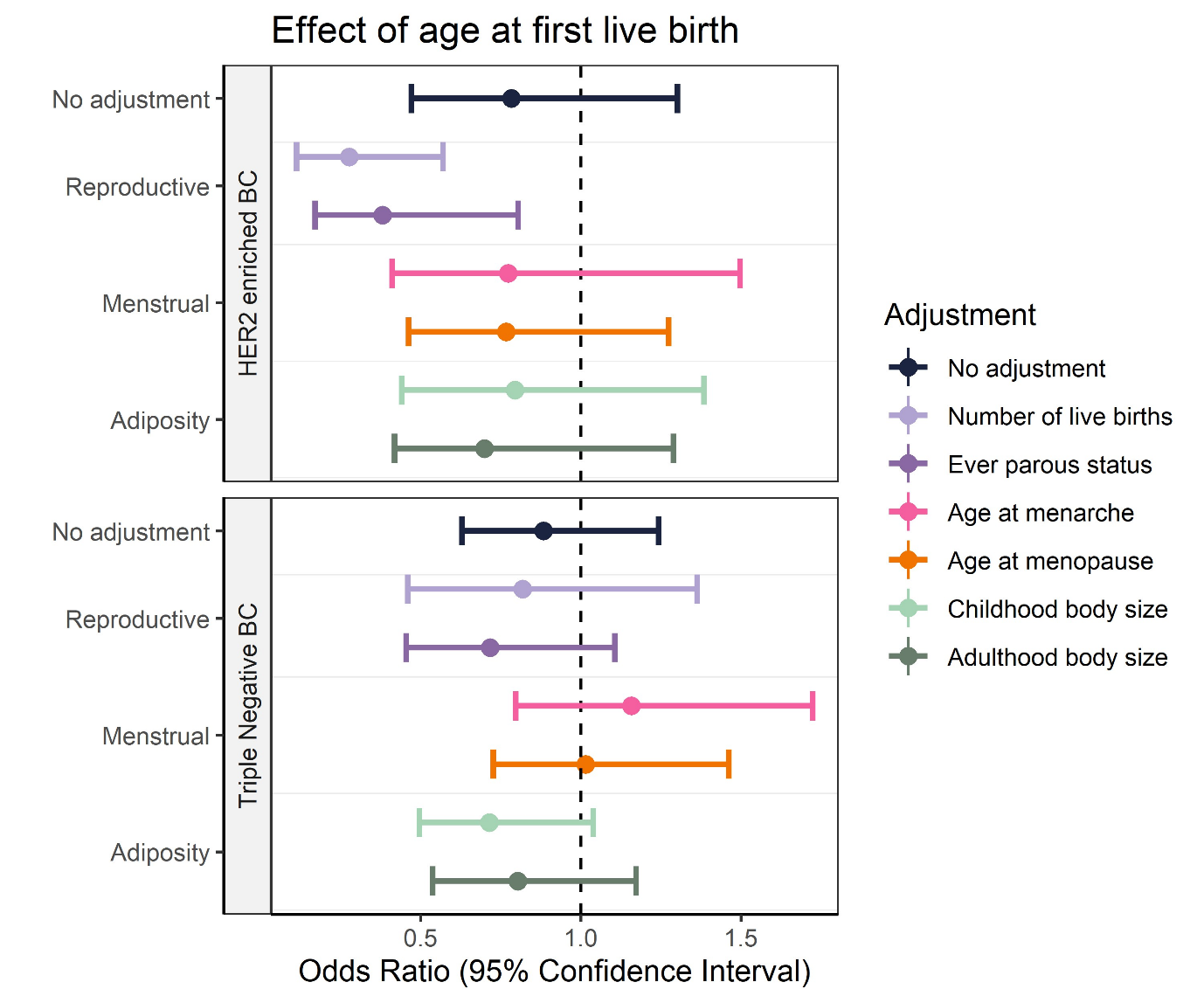
